# Dissociable Thalamocortical Circuit Disruptions During Contextual Fear Renewal in PTSD

**DOI:** 10.64898/2026.06.15.26355743

**Authors:** Muhammad Badarnee, B. Isabel Moallem, Israel Liberzon, Mohammed R. Milad

## Abstract

**Objective:** Post-traumatic stress disorder (PTSD) is marked by impaired contextual modulation of fear, leaving patients vulnerable to symptom return after extinction-based therapy. The thalamus is theorized to coordinate hippocampal-prefrontal circuits during contextual updating. Yet, its role in PTSD remains uncharacterized. We examined the contribution of the medial mediodorsal thalamus (MDm) to extinction-recall and fear renewal and its association with symptom severity.

**Methods:** 425 participants completed threat renewal and 524 extinction-recall paradigms during fMRI (threat renewal: 189 healthy controls, HC; 129 trauma-exposed HC, TEHC; 107 PTSD extinction-recall: 280 HC; 132 TEHC; 112 PTSD). Analyses examined MDm activation and connectivity with canonical fear-regions; lateral mediodorsal thalamus (MDl) and anterior pulvinar served as control regions. Structural equation modeling characterized the covariance linking thalamocortical connectivity to diagnostic group.

**Results:** During fear renewal but not extinction-recall, a Time × Group interaction emerged in MDm functional connectivity: PTSD participants showed reduced MDm connectivity with hippocampus and sgACC relative to control groups during early but not late fear renewal. Parallel reductions emerged in anterior pulvinar-vmPFC connectivity. MDl, showed no group differences. Structural equation modeling indicated that thalamo-hippocampal connectivity covaried with group via both MDm-sgACC and anterior pulvinar-vmPFC connectivity. MDm-dACC connectivity scaled with PTSD severity, independent of MDl and anterior pulvinar

**Conclusions:** State-specific reductions in MDm-hippocampal-cingulate and pulvinar-vmPFC connectivity during early fear renewal in PTSD highlight parallel thalamocortical alterations during flexible contextual threat updating. These alterations, along with the selective MDm-dACC association with symptom severity, nominate MDm-centered circuit as a hypothesis-generating focus for future mechanistic neuromodulation studies.

## Introduction

The hallmark of successful post-traumatic stress disorder (PTSD) treatment is the ability to generalize safety learning across diverse environments. However, the ‘fear renewal effect’ – the context-dependent recovery of extinguished fear – remains the primary obstacle to long-term clinical recovery, explaining why many patients relapse outside of the therapeutic setting. Current neurobiological models propose that this vulnerability stems from fundamental challenges in contextual processing, where the brain fails to appropriately gate inhibitory safety signals based on environmental cues(1–3). While substantial research has focused on the hippocampal-medial prefrontal cortex axis (mPFC), the thalamus remains comparatively underexplored. In particular, the mediodorsal thalamus (MD) which orchestrates prefrontal circuits through dense, reciprocal connections with the PFC. Rodent tracing studies have long defined PFC subregions by their MD inputs, highlighting this nucleus’ role in shaping prefrontal computations(4–7). Through these pathways, the MD supports behavioral adapting to changing threat contingencies(8–10). Anatomically, the medial subdivision of the MD (MDm) is uniquely positioned to integrate prefrontal and limbic signals through reciprocal projections with the mPFC and the amygdala(4, 11–14). This connectivity profile places the MDm at the intersection of affective salience and cognitive appraisal of threat value. Yet, whether MDm-PFC-hippocampus interactions facilitate adaptive threat assessment or contribute to maladaptive responses in PTSD remains unknown.

Causal manipulations in rodent models demonstrate that the MD is necessary for sustaining prefrontal representations that support threat detection and memory updating(15–17). Early lesion studies showed that MD damage slows fear acquisition in rabbits and attenuates conditioned freezing in rats when performed either before or after Pavlovian conditioning, indicating that the MD is required for both the formation and expression of conditioned fear memories(18, 19). The mechanistic account of these findings has been refined by a recent study demonstrating that extinction learning depends on the firing mode of MD neurons. During extinction, wild-type mice exhibit a selective increase in tonic firing within the MD, a pattern absent in mutant mice; critically, stimulation of this structure rescues extinction deficits(15). Complementing these findings, electrical stimulation of the MD during extinction reliably evokes field potentials in the medial PFC and induces state-dependent plasticity in thalamo-prefrontal transmission(20). Together, these rodent studies demonstrate that the MD causally regulates prefrontal plasticity and firing dynamics required for adaptive modification of threat processing.

In humans, large-scale meta-analytic evidence implicates the thalamus and the MD in particular in fear conditioning(21). We recently identified MD activation signals consistent with threat learning and salience processing in healthy individuals(8). In clinical populations, overall functional and structural thalamic alterations have been reported, particularly in PTSD(22, 23). Resting-state investigations demonstrate disrupted thalamo-cortical connectivity in PTSD, with abnormalities associated with symptom severity and clinical outcomes(24–26). Reductions in thalamic volume have been observed in higher-order nuclei, including the MD(27, 28).

Yet, the circuit mechanisms through which higher-order thalamic nuclei influence contextual threat processing in humans remain unclear. Addressing this gap is critical for understanding how abnormalities in thalamic circuitry may promote context-dependent reinstatement of fear. In this cross-sectional study we examined MDm activation and connectivity with canonical fear-related regions, with hypotheses formulated a priori regarding altered connectivity during contextual updating of fear. This focus reflects the MDm’s dense connectivity with limbic and mPFC regions(4, 11–14). To evaluate anatomical specificity, we included the lateral MD (MDl) and anterior pulvinar as control regions.

We recently proposed the anterior pulvinar and MD as distinct mediators of “low- and high-road” fear pathways, respectively(8), consistent with dual-route models of fear processing(29). Here, we hypothesized that clinical symptom severity would uniquely track MDm connectivity rather than the anterior pulvinar. Specifically, given that the rodent prelimbic cortex (human dACC homolog) promotes fear expression while the infralimbic cortex (human vmPFC/sgACC homologs) inhibits fear(3, 30), we predicted that greater PTSD symptom severity would map onto MDm connectivity with key fear-expression nodes (dACC, amygdala, and hippocampus). To ensure these associations were specific to fear-expression circuitry, our analyses controlled for MDm connectivity with inhibitory regions (vmPFC and sgACC).

## Methods

### Participants

We analyzed fMRI data during fear renewal (N = 425: 189 healthy controls [HC], 129 trauma-exposed healthy controls, 107 PTSD) and extinction recall (N = 524: 280 HC, 132 TEHC, 112 PTSD); See demographic characteristics in **Supplementary Tables S1-S2**. Participants were right-handed, English-speaking adults (aged 18–67 years; both sexes) with normal or corrected-to-normal vision who met standard MRI exclusion criteria. Current PTSD diagnosis and symptom severity were assessed using the Clinician-Administered PTSD Scale (see **Supplementary Information** for comprehensive clinical and demographic details).

This study was conducted in accordance with the Declaration of Helsinki and approved by the Partners HealthCare Institutional Review Board (Massachusetts General Hospital, Harvard Medical School). All participants provided written informed consent.

### Procedure

Participants underwent a two-day Pavlovian fear conditioning paradigm in the MRI (31, 32) including extinction-recall and fear renewal (**Figure 1-2A**). Retrieval occurred 24h after acquisition and extinction learning without reinforcement. Full task details are provided in the **Supplementary Information**.

**Figure 1.**
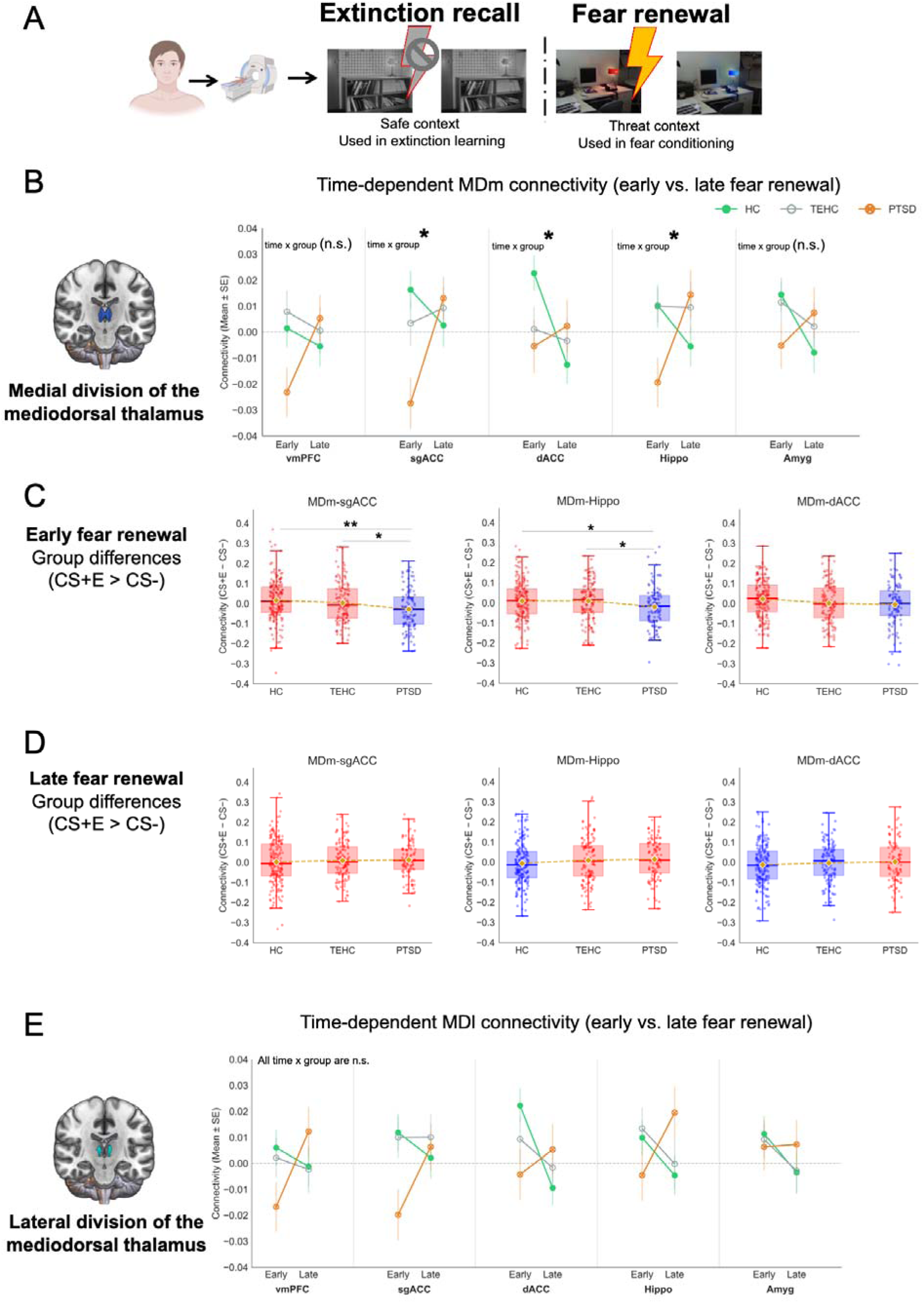
Time × Group interactions in mediodorsal thalamic connectivity with canonical fear regions during fear renewal. (A) Experimental paradigm. (B) Time × Group interaction in MDm connectivity. (C) Follow-up group comparisons during early fear renewal. (D) Follow-up group comparisons during late fear renewal. (E) Time × Group interaction in MDl connectivity. Box plots depict median and interquartile range; individual data points are shown. Significant post-hoc comparisons are indicated: *p < 0.05, **p < 0.01, FDR-corrected. *N* = 425. HC = healthy controls; TEHC = trauma-exposed healthy controls; PTSD = posttraumatic stress disorder; MDm = medial division of the mediodorsal thalamus; MDl = lateral division of the mediodorsal thalamus; vmPFC = ventromedial prefrontal cortex; dACC = dorsal anterior cingulate cortex; sgACC = subgenual anterior cingulate cortex; Hippo = Hippocampus; Amyg = Amygdala; Panel (A) was created with BioRender (https://BioRender.com/f7dzdic)

**Figure 2.**
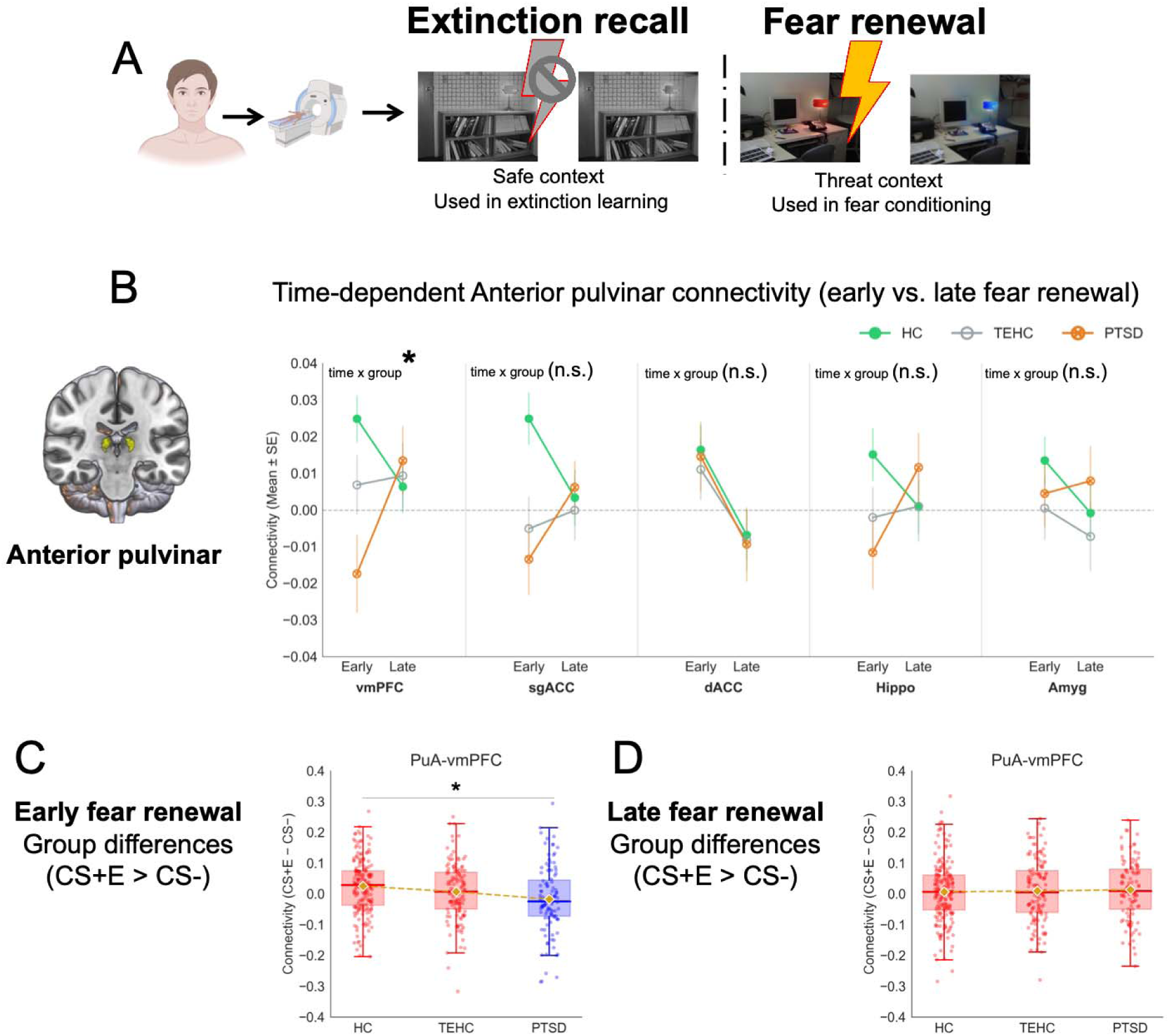
Time × Group interactions in anterior pulvinar connectivity with canonical fear regions during fear renewal. (A) Experimental paradigm. (B) Time × Group interaction. (C) Follow-up group comparisons during early fear renewal. (D) Follow-up group comparisons during late fear renewal. Box plots depict median and interquartile range; individual data points are shown. Significant post-hoc comparisons are indicated: *p < 0.05, FDR-corrected. *N* = 425. HC = healthy controls; TEHC = trauma-exposed healthy controls; PTSD = posttraumatic stress disorder; MDm = medial division of the mediodorsal thalamus; MDl = lateral division of the mediodorsal thalamus; vmPFC = ventromedial prefrontal cortex; dACC = dorsal anterior cingulate cortex; sgACC = subgenual anterior cingulate cortex; Hippo = Hippocampus; Amyg = Amygdala; Panel (A) was created with BioRender (https://BioRender.com/f7dzdic)

### Data Acquisition and Analyses

fMRI data were acquired on a 3T scanner and preprocessed using fMRIPrep (v20.0.2). Regions of interest were defined using the Automated Anatomical Labelling Atlas(33) and Neurosynth-derived coordinates(34) (see **Supplementary Information**). Activation (CS+E > CS−) was estimated using SPM12, and thalamic connectivity using gPPI in CONN with seeds (MDm, MDl, anterior pulvinar) and targets (vmPFC, sgACC, dACC, and anatomically defined amygdala, and hippocampus). Details on preprocessing and models are provided in the **Supplementary Information**.

For each phase, early and late windows were defined as the first-4 and last-4 trials, respectively. Statistical analyses included mixed RM-ANOVA, correlations, regression models, and structural equation modeling; correction for multiple comparisons was applied where applicable (see **Supplementary Information**).

## Results

### Connectivity Alterations Specific to MDm During Fear Renewal

Using generalized psychophysiological interaction analyses, we examined CS+E > CS− connectivity between thalamic subnuclei and canonical threat-processing regions (vmPFC, sgACC, dACC, amygdala, hippocampus).

Significant Group × Time interactions in MDm connectivity during fear renewal emerged for sgACC (F_(2,422)_ = 5.18, p_FDR_ = 0.03, ηp² = 0.024), dACC (F_(2,422)_ = 3.72, p_FDR_ = 0.04, ηp² = 0.017), and hippocampus (F_(2,422)_ = 4.04, p_FDR_ = 0.04, ηp² = 0.019) but not for vmPFC and amygdala (p_FDR_ ≥ 0.10). Follow-up analyses indicated that these interactions reflected group differences in CS+E vs. CS-connectivity during early, but not late, fear renewal. During early fear renewal, MDm-sgACC connectivity differed across groups (F_(2,422)_ = 6.37, p_FDR_ = 0.005, ηp² = 0.029), explained by reduced connectivity in PTSD relative to HC (t_(294)_ = 3.55, p_FDR_ = 0.001, d = 0.43) and TEHC (t_(234)_ = 2.36, p_FDR_ = 0.02, d = 0.31), with no difference between HC and TEHC (p_FDR_ = 0.29). Similarly, MDm-hippocampal connectivity differed across groups during early fear renewal (F_(2,422)_ = 3.77, p_FDR_ = 0.03, ηp² = 0.018), reflecting reduced connectivity in PTSD relative to HC (t_(294)_ = 2.58, p_FDR_ = 0.03, d = 0.31) and TEHC (t_(234)_ = 2.25, p_FDR_ = 0.03, d = 0.30), with no HC-TEHC differences (p_FDR_ = 0.88). MDm-dACC connectivity also differed across groups during early fear renewal (F_(2,422)_ = 3.35, p_FDR_ = 0.03, ηp² = 0.016), although pairwise comparisons did not survive correction (p_FDR_ ≥ 0.054). **Figure 1**.

Activation results are reported in the **Supplementary Information** and showed no significant Group × Time interaction (**Supplementary Figure S1; Table S3**).

### Control Analyses: MDl and Anterior Pulvinar Connectivity During Fear Renewal

MDl connectivity did not show significant Group × Time interactions after FDR correction for any target region, including vmPFC (F_(2,422)_ = 2.65, p_FDR_ = 0.10, ηp² = 0.012), sgACC (F_(2,422)_ = 2.52, p_FDR_ = 0.10, ηp² = 0.012), dACC (F_(2,422)_ = 3.13, p_FDR_ = 0.10, ηp² = 0.015), amygdala (F_(2,422)_ = 0.45, p_FDR_ = 0.63, ηp² = 0.002), or hippocampus (F_(2,422)_ = 2.81, p_FDR_ = 0.10, ηp² = 0.013) (**Figure 1**; **Supplementary Table S4**), supporting the relative specificity of MDm connectivity alterations during fear renewal.

Interestingly, the anterior pulvinar–vmPFC connectivity showed a significant Group × Time interaction during fear renewal (F_(2,422)_ = 4.73, p_FDR_ = 0.04, ηp²=0.022), explained by reduced connectivity in PTSD relative to HC during early fear renewal (t_(294)_ = 3.64, p_FDR_ = 0.0009, d = 0.44). No PTSD-TEHC or HC-TEHC differences (p_FDR_ = 0.057, 0.12, respectively) **Figure 2**. No significant interactions were found for anterior pulvinar connectivity with sgACC, dACC, amygdala, or hippocampus (all p_FDR_ ≥ 0.08; **Figure 2**). Activation results are reported in the **Supplementary Information** and showed no significant Group × Time interaction during fear renewal. **Table S3, Figure S1**.

### Structural Equation Modeling of Thalamo-Hippocampal and Prefrontal Regulatory Connectivity During Early Fear Renewal

Guided by RM-ANOVA findings identifying differential connectivity across early fear renewal, we specified a structural equation model (SEM) to examine whether thalamo-hippocampal connectivity was associated with regulatory thalamo-prefrontal connectivity patterns that differentiate groups. Specifically, the model tested whether MDm-hippocampal functional connectivity was associated with MDm-sgACC and PuA-vmPFC, selected a priori based on their observed group sensitivity and their proposed involvement in infralimbic-like regulatory processes. MDm-dACC connectivity was included as a specificity control because it showed temporal modulation but did not differentiate groups. We hypothesized that stronger thalamo-hippocampal coordination during early fear renewal would be associated with stronger recruitment of regulatory thalamo-prefrontal connectivity, which in turn would be associated with group-like status. We tested this hypothesis using structural equation modeling (path analysis) with the robust weighted least-squares estimator (WLSMV), appropriate for the ordered categorical nature of the group outcome. Full model specifications are in **Supplementary Information**.

The final SEM model provided a good fit to the data, χ^2^ _(4)_ = 6.47, *p* = 0.16; CFI = 0.96; TLI = 0.94; RMSEA = 0.038; SRMR = 0.031 (**Table S6**). MDm-Hippocampus connectivity was positively associated with both MDm-sgACC (β = 0.24, *p* < 0.001, 95% CI [0.15, 0.33]) and PuA-vmPFC connectivity (β = 0.19, *p* < 0.001, 95% CI [0.09, 0.29]), but was unrelated to MDm-dACC connectivity (β = 0.008, *p* = 0.85, 95% CI [−0.08, 0.09]). Greater MDm-sgACC connectivity (β = −0.16, *p* = 0.002, 95% CI [−0.26, −0.06]) and greater PuA-vmPFC connectivity (β = −0.18, *p* < 0.001, 95% CI [−0.27, −0.08]) each associated with lower values on the group ordinal i.e., the healthy end of the spectrum (directionality of influence cannot be inferred from this cross-sectional model) (**Figure 3; Tables S7-S8**).

**Figure 3.**
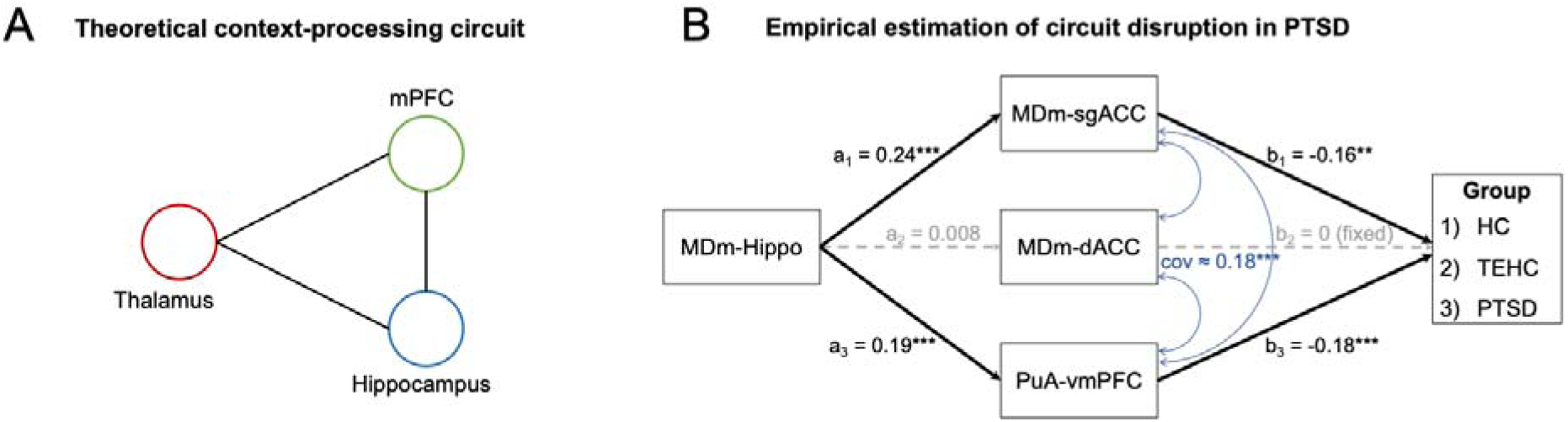
Theoretical and empirical thalamocortical circuit models of contextual fear processing in PTSD. (A) Theoretical context-processing circuit illustrating hypothesized thalamo-hippocampal-prefrontal pathways underlying contextual fear regulation. (B) Structural equation model (path analysis) empirically estimating thalamocortical circuit organization during early fear renewal. MDm-hippocampal connectivity was specified as the predictor, with MDm-sgACC, MDm-dACC, and anterior pulvinar-vmPFC connectivity as parallel mediators, and diagnostic group as the outcome. The MDm-dACC➔group path was fixed to zero as a specificity control. The model demonstrated good fit: χ²(4) = 6.47, p = 0.16; CFI = 0.96; TLI = 0.94; RMSEA = 0.038; SRMR = 0.031. Solid arrows denote significant paths; dashed arrows denote non-significant or constrained paths. Standardized coefficients are shown. **p < 0.01, ***p < 0.001. *N* = 425. MDm = medial division of the mediodorsal thalamus; PuA = anterior pulvinar; sgACC = subgenual anterior cingulate cortex; dACC = dorsal anterior cingulate cortex; vmPFC = ventromedial prefrontal cortex; PTSD = posttraumatic stress disorder; HC = healthy controls; TEHC = trauma-exposed healthy controls; SEM = structural equation modeling.

Both indirect associations were statistically significant. The effect of MDm-Hippocampus on group through MDm-sgACC connectivity was significant (β = −0.039; 95% CI [−0.068, −0.009]; p = 0.009), as was the effect through PuA-vmPFC connectivity (β = −0.034; 95% CI [−0.06, −0.008]; *p* = 0.010). The total indirect effect was significant (β = −0.073; 95% CI [−0.11, −0.033]; *p* = 0.0003).

### Specific Associations Between MDm Connectivity and PTSD Symptom Severity

To test our prior hypothesis that MDm connectivity with fear-expression nodes during fear renewal are scaled with symptom severity, we computed partial correlations between symptom severity and MDm connectivity with the dACC, amygdala, and hippocampus, each controlling for MDm connectivity with the inhibition-related nodes (vmPFC, sgACC), FDR-corrected across the three tests. During fear renewal, of the three regions tested only MDm-dACC connectivity was significantly associated with CAPS scores (r_p_ = 0.238, p_FDR_ = 0.04). This association remained significant in bootstrap analysis (10,000 resamples; 95% BCa CI [0.047, 0.40]) and permutation testing (10,000 permutations; p = 0.01; **Figure 4A, upper panel**). No significant associations were observed for MDm-amygdala (r_p_ = −0.120, p_FDR_ = 0.26) or MDm-hippocampus connectivity (r_p_ = −0.144, p_FDR_ = 0.26). **Figure 4A, lower panel.**

**Figure 4.**
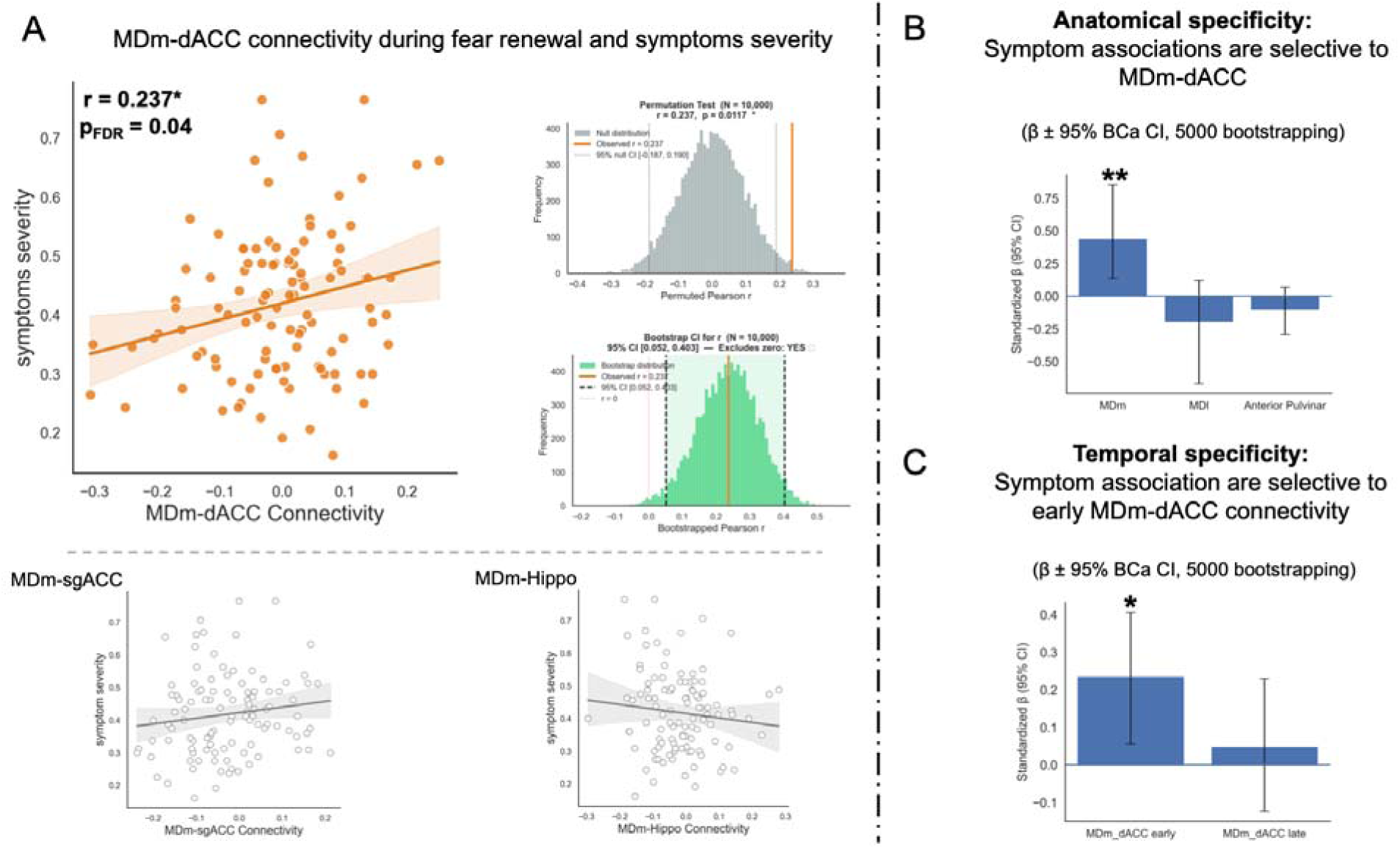
Thalamocortical connectivity and PTSD symptom severity. (A) Partial correlations between MDm-dACC, MDm-amygdala, and MDm-hippocampus and symptom severity during early fear renewal (while controlling for MDm-vmPFC, and MDm-sgACC). Only MDm-dACC connectivity was significantly associated with symptom severity. Robustness was confirmed via permutation testing (*N* = 10,000 permutations) and bootstrapping (*N* = 10,000 samples). (B) Anatomical specificity of the MDm-dACC association with symptom severity. Multiple regression including MDm-dACC, MDl-dACC, and anterior pulvinar-dACC connectivity as simultaneous predictors revealed a significant association for MDm-dACC only, confirming regional specificity. Bias-corrected and accelerated (BCa) confidence intervals were derived from 5,000 bootstrap samples. (C) Temporal specificity of the MDm-dACC association with symptom severity. Multiple regression entering early and late fear renewal MDm-dACC connectivity simultaneously revealed a significant association during early but not late fear renewal, confirming temporal specificity. BCa confidence intervals were derived from 5,000 bootstrap samples. *N* = 107 (PTSD group). dACC = dorsal anterior cingulate cortex; MDm = medial division of the mediodorsal thalamus; MDl = lateral division of the mediodorsal thalamus; sgACC = subgenual anterior cingulate cortex; PTSD = posttraumatic stress disorder.

To further assess regional specificity, and the uniqueness of MDm-dACC and symptom severity relationships, we entered the control regions (MDl-dACC and anterior pulvinar-dACC) simultaneously into a multiple regression model. MDm predicted symptoms severity (β = 0.44, p = 0.006, 95% CI [0.13, 0.87]), whereas MDl (β = −0.19, p = 0.22, 95% CI [−0.67, 0.12]) and anterior pulvinar (β = −0.11, p = 0.30, 95% CI [−0.28, 0.07]) did not (**Figure 4B**). Collinearity was acceptable (Tolerance = 0.33-0.78; VIF = 1.27-2.99), indicating nonsignificant associations were not attributable to multicollinearity. Together, symptom severity is selectively scaled with MDm rather than reflecting shared variance across thalamic nuclei.

Finally, to assess temporal specificity, MDm-dACC connectivity during early and late fear renewal were entered simultaneously into a multiple regression model predicting CAPS scores. Only early MDm-dACC connectivity remained significantly associated with CAPS scores (β = 0.23, p = 0.015, 95% CI [0.059, 0.40]), while the late was not significant (β = 0.04, p = 0.61, 95% CI [−0.12, 0.23]), **Figure 4C**. Collinearity was acceptable (Tolerance = 0.99; VIF = 1.003), indicating nonsignificant associations were not attributable to multicollinearity. Together, PTSD symptom severity is selectively associated with MDm rather than reflecting shared variance across thalamic nuclei.

### Extinction Recall

Extinction recall analyses showed no significant Group × Time interactions for either connectivity or task-related activation across MDm, MDl, or anterior pulvinar (all p_FDR_ > 0.05; **Supplementary Figure S2; Table S3-S4**).

## Discussion

These findings provide empirical support for dissociable thalamocortical alterations during contextual fear renewal in PTSD, extending a theoretically proposed hippocampal-thalamo-mPFC circuit by characterizing its disruption with anatomical (MDm) and temporal (early fear renewal) specificity. Although MDm activation was preserved, early fear renewal was marked by reduced MDm functional connectivity with hippocampal and cingulate regions, suggesting impaired thalamocortical integration rather than focal regional dysfunction. These alterations differentiated PTSD from control groups across MDm-hippocampal and MDm-sgACC connectivity, whereas MDm-dACC connectivity scaled with symptom severity within the PTSD group. Unexpectedly anterior pulvinar-vmPFC connectivity also differentiated groups, potentially pointing to parallel disruptions across thalamocortical pathways. Critically, these alterations were anatomically specific to the MDm and anterior pulvinar – nuclei previously proposed as thalamic substrates mediating cortically guided and subcortical threat processing, respectively(8). The distinct reduced connectivity findings observed in each nucleus provide the first empirical support of these hypothesized roles during fear renewal in PTSD.

It has been proposed that PTSD is associated with impairments in contextual processing rather than primary dysfunctions in threat acquisition or extinction learning per se(31). Contemporary models further suggest that these deficits arise from dysfunction within a hippocampal-thalamo-mPFC circuit(1). The hippocampal-MDm-cingulate disruptions findings observed in this study extend this framework by identifying anatomically and temporally specific abnormalities during early fear renewal. The significant time × group interaction, arising from group differences during early but not late fear renewal (PTSD vs. HC and TEHC), refines the functional interpretation of MDm-centered circuitry. Early fear renewal constitutes a phase of contextual competition, wherein the CS+E simultaneously evokes threat and extinction representations while the surrounding context reinstates prior threat associations(3). Adaptive responding in this phase requires efficient integration of contextual information to resolve competing predictions(3). The reduced hippocampal-MDm-cingulate connectivity observed in PTSD specifically during early fear renewal is consistent with prior evidence that PTSD is associated with a failure to use contextual information (35), and may indicate reduced engagement of this circuit during the contextual integration involved in fear renewal.

Extending beyond our primary hypothesis, anterior pulvinar-vmPFC connectivity differentiated groups during early fear renewal, suggesting that PTSD-related disruptions involve pulvinar-prefrontal regulatory circuitry in addition to hippocampal-MDm-cingulate pathways. SEM findings revealed parallel indirect pathways whereby stronger thalamo-hippocampal connectivity was associated with both MDm-sgACC and anterior pulvinar-vmPFC connectivity; patterns characteristic of the healthy end of the diagnostic spectrum, whereas weaker regulatory connectivity was associated with patient-like status. Within contextual accounts of fear renewal, these findings suggest that thalamo-hippocampal connectivity covaries with distributed thalamocortical regulatory connectivity during updating the contextual cues of extinguished threat. The absence of mediation through MDm-dACC further suggests architectural specificity within this thalamic circuit. Although speculative, early fear renewal might reflect coordinated interactions between contextual and hippocampal-thalamocortical regulatory processes, a pattern absent during late fear renewal when associative contingencies have stabilized and contextual integration demands are reduced – consistent with MDm-centered circuitry being preferentially engaged during competition-driven stages of fear updating.

The MDm-dACC connectivity association with symptom severity is temporally and anatomically selective, emerging uniquely during early fear renewal and absent in other thalamic regions. However, this association may appear counterintuitive, as greater coupling would be expected to reflect more normative circuit function. However, this pattern can be interpreted within established models of prefrontal-amygdala regulation. It has been proposed that prelimbic regions promote fear expression, whereas infralimbic regions support fear inhibition(3). Translational accounts suggest that these functions map onto dACC and vmPFC, respectively, in humans(30). In this framework, heightened MDm-dACC connectivity in PTSD may reflect increased engagement of a fear-expression system in the context of impaired vmPFC-mediated regulation, which is has been reported in PTSD(31, 36). This interpretation is consistent with the suggestion that low vmPFC/dACC activation ratio tips the balance towards more fear expression in PTSD(37).

Despite evidence of impaired recall performance in PTSD(31), we did not observe group differences in MDm-centered connectivity during extinction recall. Both extinction recall and fear renewal involve re-presentation of the same CS+E and differ only in context. Thus, the MDm is likely engaged across both phases. The divergence between the phases reflects how contextual information modulates the circuit rather than selective recruitment of MDm by threat. Accordingly, our findings indicate that PTSD-related alterations in hippocampal-MDm-prefrontal connectivity emerge preferentially when the context signals threat, during early fear renewal, rather than during retrieval of extinction memory in a safe context.

### Clinical Directions

Fear relapse remains a central clinical challenge in PTSD, as symptoms frequently re-emerge following interventions. Although trauma-focused therapies such as cognitive processing therapy and prolonged exposure are efficacious, a substantial proportion of patients continue to meet diagnostic criteria following treatment, and many others fail to achieve clinically meaningful improvement. It has been estimated that non-response varies widely across studies, with approximately 20-67% of patients showing limited benefit from exposure-based interventions(38). Additionally, a review paper examining clinical trials of PTSD therapy, reported that around two-thirds of the patients retained the PTSD diagnoses after treatment(38). Moreover, approximately 36% of those who showed reliable recovery in therapy, reported symptom relapse after therapy(39). Together, these results reflect heterogeneity in treatment response and difficulty in sustaining recovery after completing PTSD treatment.

The reduced MDm-centered connectivity we observed during fear renewal may be relevant to symptom persistence and relapse in PTSD for two possible reasons. First, this circuit is suggested to support using context to set the adaptive fear response. When this circuit is under engaged, fear may become poorly matched to context, either returning where it is no longer warranted or failing to track genuine changes in threat. Second, reduced engagement of this circuit may reflect diminished sensitivity to contextual threat cues that ordinarily serve adaptive, protective functions. If contextual fear is not appropriately mobilized when threat is present, this could contribute to re-exposure to hazardous situations or to the accumulation of subsequent stressors. This is consistent with proposals that impaired contextual processing in PTSD can manifest as a failure to identify genuine danger(35). These interpretations are speculative and were not directly tested, as we did not collect treatment or behavioral data

Advances in neuromodulation technologies, including focused ultrasound and other approaches capable of targeting deep brain structures, provide a potential avenue for testing the clinical implication of the MDm-centered thalamocortical circuitry. Within this framework, MDm-dACC coupling during early fear renewal is a candidate circuit for future interventional research. Notably, the anatomical specificity of the MDm offers a temporally precise target during periods of stimulus ambiguity in a threat context, such as early fear renewal. Establishing this circuit as a treatment target would require longitudinal and causal (e.g., neuromodulation) evidence not provided here, and non-invasive modulation of a structure as small and deep as the MDm remains technically demanding. Whether combining such circuit-targeted approaches with conventional therapies reduces relapse risk is an open empirical question.

Thus, future studies could therefore evaluate whether targeted modulation of this hippocampal-MDm-cingulate pathway alters early fear renewal processes and enhances the durability of exposure-based treatments in individuals with PTSD. Such work will be essential to determine whether interventions directed at this thalamocortical control circuit can produce measurable and clinically meaningful improvements in symptom persistence and relapse risk.

### Limitations

We acknowledge several limitations to the present study. Finer parcellation of the MDm and the control regions would offer greater anatomical specificity. Additionally, fMRI lacks the resolution to distinguish between different neuronal populations within the thalamic nuclei. As such, our conclusions are constrained to the aggregate BOLD signal of each region of interest and cannot capture the cell-level dynamics or neuroplasticity. Given the inherent limitations of human neuroimaging, this study’s correlational design precludes causal inference. Interventional studies that directly manipulate specific brain regions such as the MDm and assess their downstream effects are essential to establish causality. Although the observed effect sizes are modest by conventional statistical standards, such magnitudes are typical in neuroimaging tools and clinical measures(40). These effects were stable across robustness checks but small in magnitude; their clinical relevance remains to be established in studies that link connectivity to longitudinal, patient-reported outcomes. Finally, our samples were predominantly composed of White and Asian participants; consequently, these findings may not fully generalize to other racially and ethnically diverse populations.

## Data Availability

All data produced in the present study are available upon reasonable request to the corresponding author.

## Disclosures

All authors report no competing interests.

## Acknowledgments

This study has been supported by the National Institute of Mental Health to Dr. Milad: R01MH123736, R01MH125198, R33MH111907, R01MH097880, and R01MH097964

## Notes

### Competing Interest Statement

The authors have declared no competing interest.

### Author Declarations

This study was conducted in accordance with the Declaration of Helsinki and approved by the Partners HealthCare Institutional Review Board (Massachusetts General Hospital, Harvard Medical School).

