## Supplementary Information for "Dissociable Thalamocortical Circuit Disruptions During Contextual Fear Renewal in PTSD"

### Supplementary Methods

#### Participants

The demographic characteristics of the study sample is presented in **Tables S1-S2**. The exclusion criteria were consistent with standard MRI studies and included significant head trauma, history of seizures, current substance abuse or dependence, metal implants, pregnancy, breastfeeding, and a positive urine toxicology screen. Participants with post-traumatic stress disorder (PTSD) were required to be either medication-free for at least 8 weeks or on a stable medication regimen for at least 8 weeks. PTSD diagnosis was confirmed using the Clinician-Administered PTSD Scale (CAPS). Some data from this sample have been reported previously with a different focus(1–3); the present analyses are novel.

**Supplementary Table S1.** Demographic Characteristics of the Fear Renewal Sample by Group

| **Characteristic** | **HC (n = 189)** | **TEHC (n = 129)** | **PTSD (n = 107)** |
|---|---|---|---|
| **Age, years** | | | |
| Mean ± SD | 26.7 ± 7.8 | 34.5 ± 16.4 | 32.1 ±13.7 |
| Unknown, n (%) | 29 (15.3) | 1 (0.8) | 0 (0.0) |
| **Biological sex, n (%)** | | | |
| Female | 125 (69.4) | 81 (62.8) | 78 (72.9) |
| Male | 55 (30.6) | 48 (37.2) | 29 (27.1) |
| Unknown | 9 (4.8) | 0 (0.0) | 0 (0.0) |
| **Race, n (%)** | | | |
| Asian | 26 (17.0) | 8 (7.3) | 8 (7.8) |

| | | | |
|---|---|---|---|
| White | 110 (71.9) | 75 (68.8) | 64 (62.1) |
| Black | 5 (2.6%) | 17 (13.2%) | 16 (15.5) |
| Other/Multiracial | 12 (7.8) | 9 (8.3) | 15 (14.6) |
| Unknown | 36 (19.0) | 20 (15.5) | 4 (3.7) |

**Note.** HC = healthy control; TEHC = trauma-exposed healthy control; PTSD = post-traumatic stress disorder. Percentages for sex and race are calculated among participants with available data (valid percentages); the Unknown row is reported as n (% of the full group N).

**Supplementary Table S2.** Demographic Characteristics of the Extinction Recall Sample by Group

| **Characteristic** | **HC (n = 280)** | **TEHC (n = 132)** | **PTSD (n = 112)** |
|---|---|---|---|
| **Age, years** | | | |
| Mean ± SD | 28.1 ± 8.1 | 35.2 ± 16.8 | 33.0 ±14.4 |
| Unknown, n (%) | 31 (11.1) | 4 (3.0) | 0 (0.0) |
| **Biological sex, n (%)** | | | |
| Female | 185 (68.8) | 82 (63.6) | 80 (71.4) |
| Male | 84 (31.2) | 47 (36.4) | 32 (28.6) |
| Unknown | 11 (3.9) | 3 (2.3) | 0 (0.0) |
| **Race, n (%)** | | | |
| Asian | 37 (15.4) | 8 (7.1) | 8 (7.5) |
| White | 176 (73.3) | 76 (67.9) | 66 (61.7) |

| | | | |
|---|---|---|---|
| Black | 10 (4.2) | 18 (16.1) | 18 (16.8) |
| Other/Multiracial | 17 (7.1) | 10 (8.9) | 15 (14.0) |
| Unknown | 40 (14.3) | 20 (15.2) | 5 (4.5) |

**Note.** HC = healthy control; TEHC = trauma-exposed healthy control; PTSD = post-traumatic stress disorder. Percentages for sex and race are calculated among participants with available data (valid percentages); the Unknown row is reported as n (% of the full group N).

**Experimental Paradigm**

We employed a four-phase Pavlovian fear conditioning paradigm to examine contextual threat processing during extinction recall and fear renewal(1, 4). Neutral visual stimuli (colored lights) served as conditioned stimuli (CS+ and CS-), and mild electric shocks served as unconditioned stimuli. Before conditioning, an individualized calibration procedure determined shock intensity, starting at 0.1 mA and incrementing until rated as "highly annoying but not painful," with a maximum of 20 mA for safety and ethical reasons.

On Day 1, participants underwent threat acquisition in Context A (e.g., office room), where the CS+ was partially reinforced with electric shock (62.5% reinforcement; 500 ms duration) and the CS- was never reinforced. Extinction learning was subsequently conducted in a distinct context (Context B; e.g., bookcase room), where both stimuli were presented without reinforcement.

On Day 2, all analyses were conducted. Extinction recall was tested in Context B, followed by fear renewal testing in the original acquisition context (Context A). All stimuli were presented without shock reinforcement. This design allowed examination of context-dependent fear responding. Specifically, the return of extinguished fear upon return to the acquisition context.

Across all phases, stimuli were presented for 6 s with randomized inter-trial intervals (fixation screen; 12–18 s). Trial order was pseudorandomized and counterbalanced across participants.

### Clinical Assessments

#### PTSD Symptoms

PTSD symptom severity was assessed using the Clinician-Administered PTSD Scale (CAPS) for DSM-IV or DSM-5(5). Total severity scores were computed as the sum of frequency and intensity ratings across all items. To harmonize scores across versions, each total score was converted to a proportion of the maximum possible score (CAPS-IV: total/136; CAPS-5: total/80). PTSD diagnosis was confirmed in the PTSD group using CAPS criteria; trauma-exposed healthy controls did not meet diagnostic threshold.

### MRI Data Acquisition and Preprocessing

We collected the MRI data using a Trio 3T whole-body MRI scanner (Siemens Medical Systems, Iselin, New Jersey) using an 8/32-channel head coil. The functional data in this setting were acquired using a T2* weighted echo-planar pulse sequence with these parameters: TR = 2.56/3.0s, TE = 30 ms, slice number = 45/48, voxel size = $3 \times 3 \times 3$ mm$^3$. The anatomical images were collected using a T1-weighted MP-RAGE pulse sequence, parcellated into $1 \times 1 \times 1$ mm$^3$ voxels. Across subjects, we used elastic bands on the head coil device to reduce head motions.

We preprocessed the neuroimaging data using the default pipeline in fMRIPrep, version 20.0.2 (6, 7). The process included standard steps of correction of slice timing, realignment of the functional images, and coregistration. Additionally, we normalized the data into the Montreal Neurological Institute (MNI) space and smoothed with a 6-mm full-width half-maximum Gaussian kernel.

**fMRI Activation Analyses**

We used SPM 12 to estimate the activation within the MDm and control regions, i.e., MDl and anterior pulvinar. Specifically, for each subject, we applied the least-squares-based generalized linear model (GLM) and estimated the beta values for each voxel within the regions of interest in response to CS+ vs. CS-. We included 32 regressors for the CS+ and CS-, a regressor for the context, and a regressor for the electric shock in fear learning but not in retrieval. The GLM also included the typical six head movement parameters (x, y, z directions, and rotations). We used the contrast maps resulting from the first-level analysis to estimate the variability of these maps across participants at the group-level analysis. Then, we used the contrast maps resulting from the group-level analysis to extract the averaged values across all voxels within the predefined masks of the MDm, MDl, and anterior pulvinar. We used this final output to compare the BOLD response during the first two trials of each learning phase.

**Functional Connectivity Analyses**

We used the CONN functional connectivity toolbox, version 22. a, for the MathWorks MATLAB program, to compute the connectivity values (8, 9). We started by segmenting the brain images into gray matter, white matter, and CSF. Then, we applied a standard CONN toolbox denoising pipeline (10) to the functional images to control the effect of potential confounding parameters, using a component-based noise correction method. Along with applying bandpass frequency filtering of the BOLD time series to the range of between 0.008 Hz and 0.09 Hz.

To evaluate the differences in connectivity between CS+ and CS−, we used generalized psychophysiological interaction (gPPI) analyses. We defined the first four trials of each condition as a block. The MDm and MDl were defined as individual seed regions, and connectivity was assessed with five target regions: the dACC, sgACC, vmPFC, amygdala, and hippocampus. The gPPI model included seed BOLD signals as physiological factors, boxcar signals characterizing the task conditions convolved with an SPM canonical hemodynamic response function as psychological factors, and the product of the two as psychophysiological interaction terms. Functional connectivity changes were quantified using Fisher-transformed correlation coefficients of the psychophysiological interaction terms. At the group level, we used a GLM to assess task-related connectivity changes across participants.

**Multivariate Analysis**

**Group-Level Comparisons**

Activation and connectivity during extinction retrieval and fear renewal were each segmented into early and late blocks, defined as the average response to CS+E versus CS− across four consecutive trials per block.

Group differences were examined using a 2 × 3 repeated-measures ANOVA with Time (early vs. late) and Group (HC, TEHC, PTSD) as factors. When a significant Time × Group interaction was detected, follow-up analyses examined group differences separately for early and late phases. Post-hoc pairwise comparisons were conducted with FDR correction applied within each time phase. Given our a priori hypothesis of group differences during early but not late phases, this segmentation was theoretically motivated rather than exploratory.

**Structural Equation Modeling**

**Rationale for Structural Equation Modeling**

Relations among fear-renewal functional connectivity measures and diagnostic group were examined using structural equation modeling (path analysis) rather than ordinary regression-based mediation. This choice was driven by the nature of the outcome variable: diagnostic group was an ordered categorical variable with three levels (healthy controls, trauma-exposed healthy controls, and patients with PTSD). Conventional mediation frameworks assume a continuous, normally distributed outcome with a linear link. These assumptions are not appropriate for an ordinal outcome. Instead, structural equation modeling with the robust weighted least-squares estimator (WLSMV) models are recommended when the outcome variable is ordinal. These models treat the ordered categorical outcome through an underlying latent response variable and estimated thresholds via a probit formulation. Thereby, WLSMV models accommodate the ordinal structure of the group variable.

Additionally, structural equation modeling allows estimating all paths simultaneously within a single model, modeling residual covariances among correlated mediators explicitly, defining theory-driven constraints, estimating indirect effects with standard errors and confidence intervals, and evaluating the overall model fit. These features together are not available in a regression-based mediation model.

**Model Specification**

We specified the structural equation model in two steps, each motivated by a prior theoretical and/or empirical considerations. The overall directionality of the main model was specified a priori based on the hippocampal contextual-gating framework(11). In this framework, the hippocampal processing of contextual information modulates prefrontal regulation of threat-related stimuli during fear renewal. Accordingly, we treated the thalamo-hippocampal connectivity as antecedent to thalamocortical connectivity. Since this is a cross-sectional study, the proposed order in the SEM model is motivated by a theoretical assumption rather than an established analysis. The interpretation of the results, should be consistent with this assumption, rather than direction of influence.

Two specification choices were made before fitting the model:

(a) MDm-dACC connectivity was retained as a specificity control. Given the absence of group differences in MDm-dACC connectivity in the primary ANOVA, the MDm-dACC → group path was fixed to zero, allowing this pathway to serve as a test of regional specificity rather than a substantive mediator. This constraint was determined by an independent analysis conducted prior to and separately from the SEM model.

(b) Residual covariances among the three mediators were constrained to equality. The three pairs of mediators (MDm-sgACC, MDm-dACC, and PuA-vmPFC) share thalamo-cortical structure. The MDm-sgACC and MDm-dACC arise from the same MD nucleus-cingulate regions, and in our recent work, we found parallel functional roles of the MD and anterior pulvinar in fear processing(12). The shared anatomical and functional basis of these regions, motivated our decision to equality constraint their residuals.

**Model 1 (mediation with estimated direct effect)**

MDm-hippocampus connectivity was specified as the predictor and three parallel mediators were included: MDm-sgACC, MDm-dACC, and anterior pulvinar-vmPFC connectivity. Paths from the predictor to each mediator (a paths) and from mediators to group (b paths) were estimated. Along with the direct effect of MDm-Hippocampus on group (*c'*). While both indirect effects and the total indirect effect were significant, the direct effect was non-significant ($\beta = -0.064$, SE $= 0.053$, $z = -1.21$, $p = 0.23$, 95% CI [-0.17, 0.04]; full parameter estimates in **Supplementary**

**Table S7**). This pattern is consistent with an indirect-only mediation model, characterized by significant indirect effects in the absence of a significant direct effect(13, 14).

**Model 2 (final model; full mediation)**

Consistent with the non-significant direct effect observed in Model 1 and the principle of parsimony in SEM, which favors the simplest model that adequately fits the data, the c' path was constrained to zero. Indirect-effect estimates were nearly identical across the two models ($|\Delta\beta| \leq 0.007$ for all indirect paths; $|\Delta\beta| \leq 0.012$ across all estimated paths), indicating that the constraint did not influence the mediational findings. Model 2 was retained as the final model. Full parameter estimates for Model 2 are reported in Supplementary **Table S8**, and fit indices for both models are reported in Supplementary **Table S6**.

**Software**

Neuroimaging analyses were conducted using SPM12 and the CONN functional connectivity toolbox (version 22.a). Statistical analyses were performed using JASP (versions 0.18.3–0.19.3), IBM SPSS Statistics, and R (R Foundation for Statistical Computing, Vienna, Austria).

**Regions of Interest (ROI Definition)**

We used the Automated Anatomical Labelling Atlas(15) to extract predefined masks for the anterior pulvinar, MDm, and MDl, along with the two other anatomical regions we included in the connectivity analyses, i.e., the amygdala and hippocampus. Guidelines from Neurosynth (16) combined with the keyword 'conditioning' were used to create 8mm spheres around the identified peak coordinates of the functional regions: vmPFC (MNIxyz = −2, 46, −10), sgACC (MNIxyz = 0, 26, −12), and dACC (MNIxyz = 0, 14, 28).

### Supplementary Results

#### Functional activation during extinction recall and fear renewal

No significant Group × Time interactions in thalamic activation were observed during fear renewal or extinction recall among HC, TEHC, and PTSD participants (**Figure S1-S2; Table S3**).

**Table S3.** Time × Group interactions in thalamic activation during fear renewal and extinction recall (CS+E > CS−)

| **Activation** | **Fear renewal** | | **Extinction recall** | |
|---|---|---|---|---|
| | $\mathbf{F_{(2,\ 422)}}$ | **p** | $\mathbf{F_{(2,\ 521)}}$ | **p** |
| **MDm** | 1.57 | 0.20 | 0.85 | 0.42 |
| **MDl** | 1.42 | 0.24 | 0.47 | 0.62 |
| **Anterior pulvinar** | 0.82 | 0.43 | 0.15 | 0.85 |

**Note.** Results of repeated-measures ANOVA testing Time × Group interactions in functional activation for the CS+E > CS− contrast during fear renewal and extinction recall. MDm = medial division of the mediodorsal thalamus; MDl = lateral division of the mediodorsal thalamus.

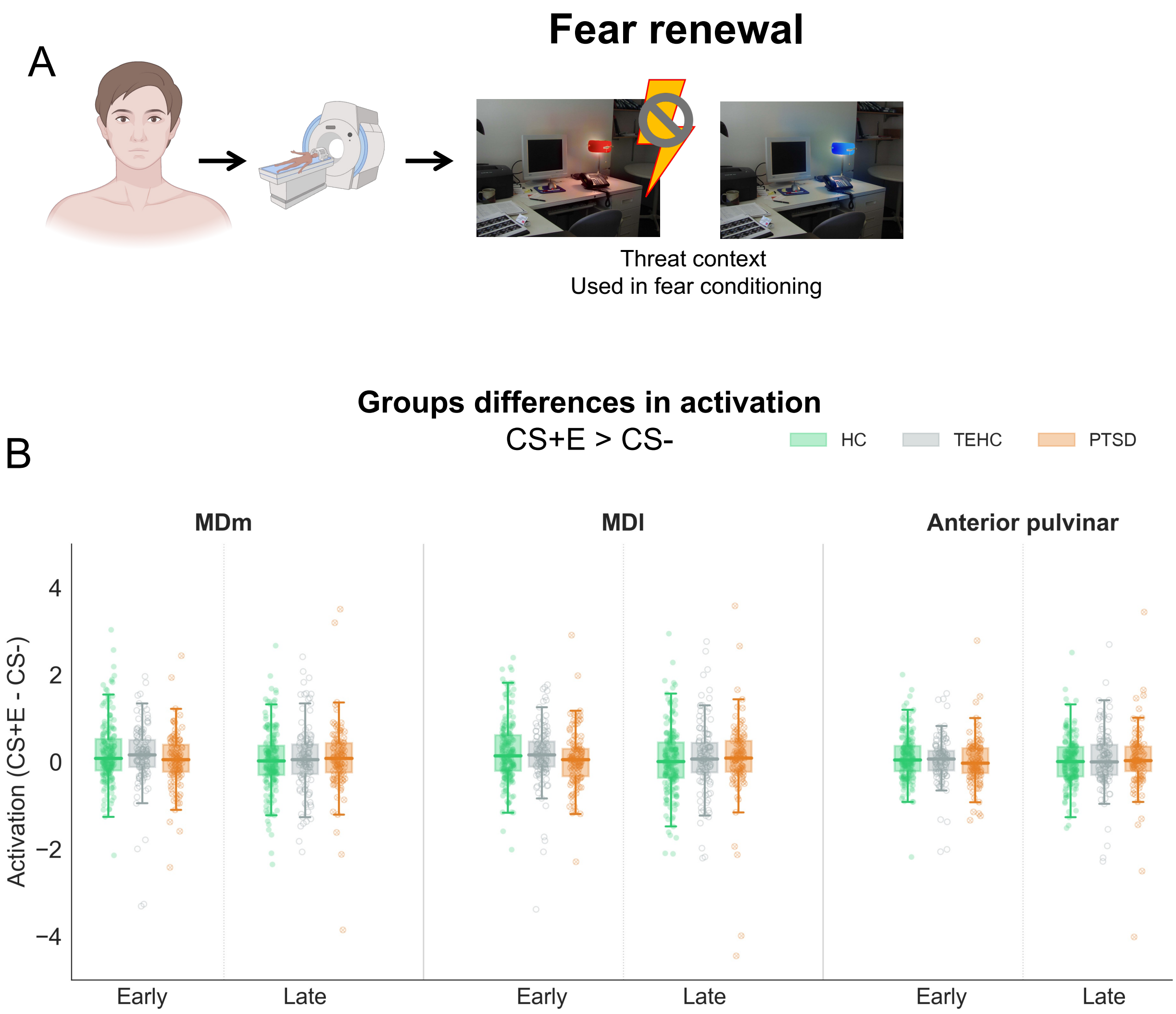


**Figure S1. Time × Group Interactions in Thalamic Activation During Fear renewal.** (A) Experimental paradigm. (B) Time × Group interaction in MDm, MDl and anterior pulvinar activation. Box plots depict median and interquartile range; individual data points are shown.

N = 425. HC = healthy controls; TEHC = trauma-exposed healthy controls; PTSD = posttraumatic stress disorder; MDm = medial division of the mediodorsal thalamus; MDl = lateral division of the mediodorsal thalamus. Panel (A) was created with BioRender (https://BioRender.com/f7dzdic)

**Functional connectivity during extinction recall**

No significant Time × Group interaction was observed in MDm connectivity with canonical fear regions (amygdala, hippocampus, sgACC, dACC, vmPFC) during extinction recall or in any thalamic control regions (**Figure S2; Table S4**).

**Table S4.** Time × Group interactions in thalamic connectivity during extinction recall (CS+E > CS−): MDm, MDl, and anterior pulvinar.

| **Connectivity** | $\mathbf{F_{(2, 521)}}$ | **p (uncorrected)** | **p (FDR)** |
|---|---|---|---|
| **Region of Interest: MDm** | | | |
| vmPFC | 0.60 | 0.55 | 0.73 |
| sgACC | 0.82 | 0.44 | 0.73 |
| dACC | 2.34 | 0.10 | 0.48 |
| Hippocampus | 0.14 | 0.87 | 0.87 |
| Amygdala | 0.54 | 0.58 | 0.73 |
| **Thalamic Control Region 1: MDl** | | | |
| vmPFC | 0.36 | 0.70 | 0.70 |
| sgACC | 0.67 | 0.51 | 0.68 |
| dACC | 2.25 | 0.11 | 0.53 |

| | | | |
|---|---|---|---|
| Hippocampus | 0.61 | 0.55 | 0.68 |
| Amygdala | 1.09 | 0.34 | 0.68 |
| **Thalamic Control Region 2: Anterior pulvinar** | | | |
| vmPFC | 0.42 | 0.66 | 0.75 |
| sgACC | 1.09 | 0.34 | 0.75 |
| dACC | 0.81 | 0.46 | 0.75 |
| Hippocampus | 0.29 | 0.75 | 0.75 |
| Amygdala | 0.76 | 0.47 | 0.75 |

**Note.** Results of repeated-measures ANOVA testing Time × Group interactions in functional connectivity for the CS+E > CS− contrast during extinction recall. Both uncorrected and FDR-corrected p-values are reported across regions. MDm = medial division of the mediodorsal thalamus; MDl = lateral division of the mediodorsal thalamus; dACC = dorsal anterior cingulate cortex; sgACC = subgenual anterior cingulate cortex; vmPFC = ventromedial prefrontal cortex.

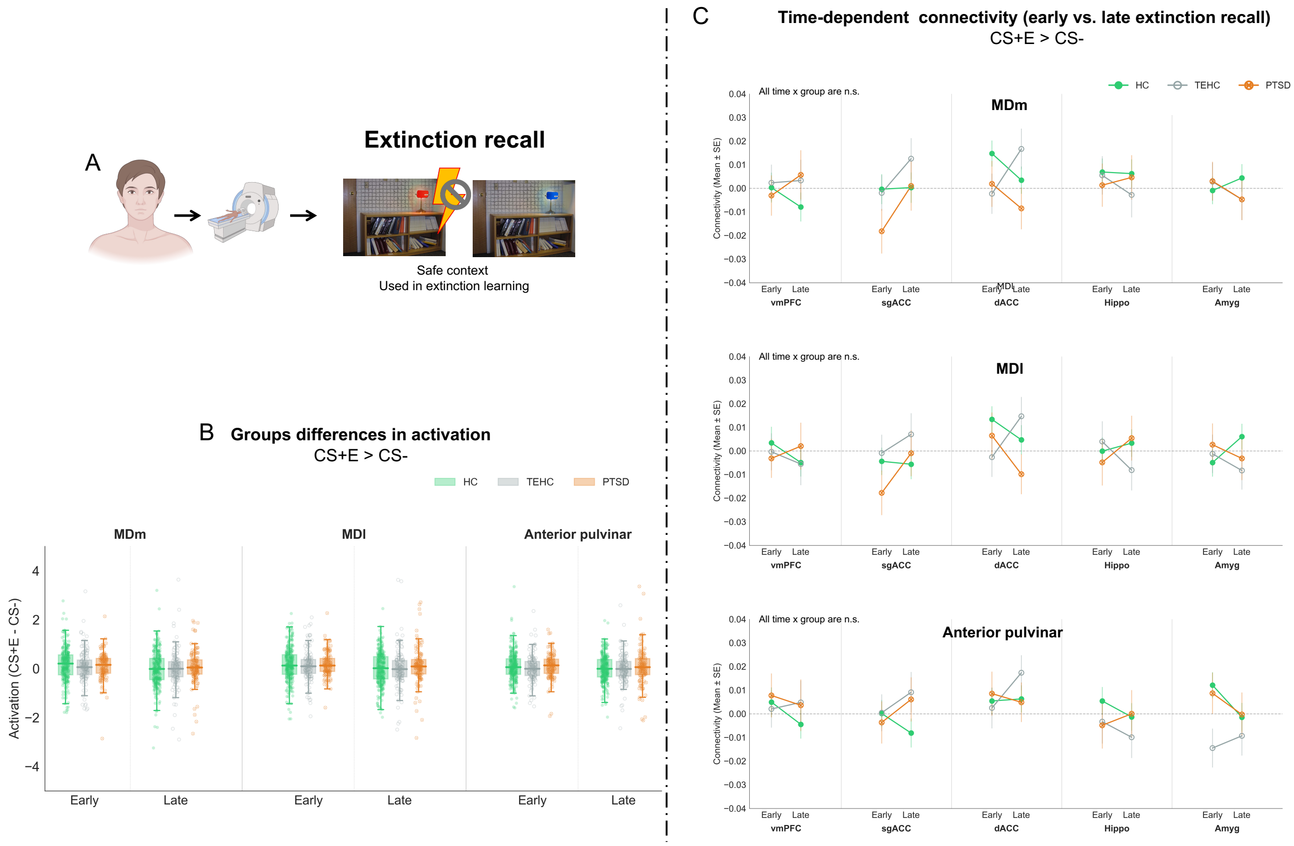


**Figure S2. Activation and Connectivity in Thalamic Nuclei of Interest During Fear renewal.** (A) Experimental paradigm. (B) Time × Group interaction in MDm, MDl and anterior pulvinar activation. Box plots depict median and interquartile range; individual data points are shown. (C) Time × Group interaction in MDm, MDl and anterior pulvinar connectivity with canonical fear regions.

N = 524. HC = healthy controls; TEHC = trauma-exposed healthy controls; PTSD = posttraumatic stress disorder; MDm = medial division of the mediodorsal thalamus; MDl = lateral division of the mediodorsal thalamus; dACC = dorsal anterior cingulate cortex; sgACC = subgenual anterior cingulate cortex; vmPFC = ventromedial prefrontal cortex. Panel (A) was created with BioRender (https://BioRender.com/7qgwsph)

**Functional connectivity during fear renewal**

Significant Time × Group interactions were observed in thalamic connectivity with canonical fear regions during fear renewal for the primary region, MDm, as well as the anterior pulvinar, whereas no significant interactions were found for the MDl (**Table S5**).

**Table S5.** Time × Group interactions in thalamic connectivity during fear renewal (CS+E > CS−): MDm, MDl, and anterior pulvinar.

| **Connectivity** | $\mathbf{F_{(2, 422)}}$ | **p (uncorrected)** | **p (FDR)** |
|---|---|---|---|
| **Region of Interest: MDm** | | | |
| vmPFC | 2.56 | 0.08 | 0.10 |
| sgACC | 5.18 | 0.006 | 0.03 |
| dACC | 3.72 | 0.02 | 0.04 |
| Hippocampus | 4.04 | 0.01 | 0.04 |
| Amygdala | 2.10 | 0.12 | 0.12 |
| **Thalamic Control Region 1: MDl** | | | |
| vmPFC | 2.65 | 0.07 | 0.10 |
| sgACC | 2.52 | 0.08 | 0.10 |
| dACC | 3.13 | 0.04 | 0.10 |

| | | | |
|---|---|---|---|
| Hippocampus | 2.81 | 0.06 | 0.10 |
| Amygdala | 0.45 | 0.64 | 0.64 |
| **Thalamic Control Region 2: Anterior pulvinar** | | | |
| vmPFC | 4.73 | 0.009 | 0.04 |
| sgACC | 3.44 | 0.03 | 0.08 |
| dACC | 0.07 | 0.93 | 0.93 |
| Hippocampus | 2.31 | 0.10 | 0.17 |
| Amygdala | 0.52 | 0.60 | 0.75 |

**Note.** Results of repeated-measures ANOVA testing Time × Group interactions in functional connectivity for the CS+E > CS− contrast during fear renewal. Both uncorrected and FDR-corrected p-values are reported across regions. MDm = medial division of the mediodorsal thalamus; MDl = lateral division of the mediodorsal thalamus; dACC = dorsal anterior cingulate cortex; sgACC = subgenual anterior cingulate cortex; vmPFC = ventromedial prefrontal cortex.

### Structural Equation Modeling

Tables S5–S7 report the structural equation modeling results described in the Supplementary Methods. Both models were estimated on N = 425. Supplementary Table S5 reports fit indices for the augmented model (Model 1; direct effect *c'* estimated) and the final model (Model 2; *c'* fixed to zero). Supplementary Tables S6 and S7 report standardized parameter estimates (β, SE, *z*, *p*, 95% CI) for Model 1 and Model 2, respectively. Both models indicated acceptable to good fit on all indices, and indirect-effect estimates were nearly identical across the two specifications.

**Table S6.** Model fit indices for the augmented (Model 1) and final (Model 2) mediation models ($N = 425$).

| Fit index | Model 1 (c' estimated) | Model 2 (final; c' = 0) |
|---|---|---|
| Sample size, N | 425 | 425 |
| Free parameters | 15 | 14 |
| $\chi^2$ | 5.037 | 6.470 |
| Degrees of freedom | 3 | 4 |
| P value, $\chi^2$ *model* | 0.169 | 0.167 |
| Comparative Fit Index (CFI) | 0.968 | 0.961 |
| Tucker–Lewis Index (TLI) | 0.936 | 0.942 |
| Bentler–Bonett Normed Fit Index (NFI) | 0.928 | 0.908 |
| Goodness of Fit Index (GFI) | 0.996 | 0.994 |
| RMSEA | 0.040 | 0.038 |
| SRMR | 0.031 | 0.031 |

**Note.** Models were estimated using robust weighted least-squares estimation (WLSMV). $\chi^2$ values represent the scaled and shifted test statistic. AIC and BIC are not available for WLSMV estimation. RMSEA = Root Mean Square Error of Approximation; SRMR = Standardized Root Mean Square Residual.

**Table S7.** Model 1: Structural equation model path estimates for thalamo-cortical-hippocampal connectivity and diagnostic group during early fear renewal, with the direct effect estimated (*N* = 425)

| Path | β | SE | Z | 95% CI | p |
|---|---|---|---|---|---|
| **Path coefficients** | | | | | |
| MDm-Hippo → MDm-sgACC | 0.24 | 0.045 | 5.19 | 0.15, 0.32 | $2.08 \times 10^{-7}$ |
| MDm-Hippo → PuA-vmPFC | 0.18 | 0.049 | 3.75 | 0.09, 0.28 | 0.0002 |
| MDm-Hippo → MDm-dACC | 0.008 | 0.044 | 0.18 | -0.08, 0.09 | 0.85 |
| MDm-sgACC → Group | -0.15 | 0.054 | -2.73 | -0.25, -0.04 | 0.006 |
| PuA-vmPFC → Group | -0.17 | 0.050 | -3.36 | -0.27, -0.07 | 0.0008 |
| MDm-dACC → Group | 0.00 | - | - | - | fixed |
| MDm-Hippo → Group | -0.064 | 0.053 | -1.21 | -0.17, 0.04 | 0.23 |
| **Indirect effects** | | | | | |
| Indirect: via MDm-sgACC | -0.035 | 0.014 | -2.40 | -0.06, -0.01 | 0.02 |
| Indirect: via PuA-vmPFC | -0.031 | 0.013 | -2.50 | -0.06, -0.01 | 0.01 |
| Total indirect effect | -0.066 | 0.019 | -3.43 | -0.10, -0.03 | 0.0006 |

**Note.** *β* = standardized coefficient; SE = standard error; CI = confidence interval estimated from the robust (expected) information matrix. The MDm-dACC → Group path was fixed to zero as a specificity control, justified by a non-significant group difference in MDm-dACC connectivity in a prior repeated-measures analysis of variance. The direct MDm-Hippo → Group path (*c'*) was estimated in this model and was non-significant; in the final model (Model 2; **Table S8**), this path was fixed to zero. Residual covariances among mediators were constrained to equality on theoretical grounds. WLSMV = weighted least squares mean and variance adjusted. Model fit indices are reported in **Table S6**.

**Table S8.** Model 2: Structural equation model path estimates for thalamo-cortical-hippocampal connectivity and diagnostic group during early fear renewal, with the direct effect constrained (N = 425)

| Path | β | SE | Z | 95% CI | p |
|---|---|---|---|---|---|
| **Path coefficients** | | | | | |
| MDm-Hippo → MDm-sgACC | 0.24 | 0.046 | 5.30 | 0.15, 0.33 | $1.14 \times 10^{-7}$ |
| MDm-Hippo → PuA-vmPFC | 0.19 | 0.051 | 3.82 | 0.09, 0.29 | 0.0001 |
| MDm-Hippo → MDm-dACC | 0.008 | 0.044 | 0.18 | -0.08, 0.09 | 0.85 |
| MDm-sgACC → Group | -0.16 | 0.053 | -3.02 | -0.26, -0.06 | 0.002 |
| PuA-vmPFC → Group | -0.18 | 0.050 | -3.52 | -0.27, -0.08 | 0.0004 |
| MDm-dACC → Group | 0.00 | - | - | - | fixed |
| **Indirect effects** | | | | | |
| Indirect: via MDm-sgACC | -0.039 | 0.015 | -2.60 | -0.07, -0.009 | 0.009 |
| Indirect: via PuA-vmPFC | -0.034 | 0.013 | -2.56 | -0.06, -0.008 | 0.01 |
| Total indirect effect | -0.073 | 0.020 | -3.61 | -0.11, -0.03 | 0.0003 |

**Note.** β = standardized coefficient; SE = standard error; CI = confidence interval estimated from robust information matrix. The dACC → group path was fixed to zero as a specificity control. The direct MDm-Hippo → group path was fixed to zero in the final model following nonsignificance in the augmented model (β = -0.064, $p = 0.23$). WLSMV = weighted least squares mean and variance adjusted. Model fit indices are reported in the main manuscript.
